# Information and genetic counselling for psychiatric risks in children with rare genomic disorders

**DOI:** 10.1101/19007294

**Authors:** Andrew Cuthbert, Aimee Challenger, Jeremy Hall, Marianne BM van den Bree

**Author notes:** Corresponding author: Marianne BM van den Bree PhD. J Hall and M van den Bree contributed equally to the work. **Contributors** A Challenger (ACh), M van den Bree (MvdB) and J Hall (JH) conceived the research question and designed the study. ACh developed the study ethics application. A Cuthbert (ACu) and ACh oversaw recruitment of study participants and data acquisition. ACu oversaw and conducted all data analysis and interpretation. ACu, JH and MvdB reviewed and discussed and revised the data analysis. ACu drafted the manuscript, figures and tables. All authors provided critical revisions to the manuscript, intellectual content and final approval for submission.

## Abstract

**Purpose:** Genomic medicine has transformed the diagnosis of neurodevelopmental disorders. Evidence of increased psychiatric comorbidity associated with genomic copy number and single nucleotide variants (CNV and SNV) may not be fully considered when providing genetic counselling. We explored parents’ experiences of genetics services and how they obtained information concerning psychiatric manifestations.

**Methods:** Parents of children diagnosed with genomic variants completed an online survey exploring, (i) how they experienced the genetic diagnosis, and (ii) how they acquired information about psychiatric, developmental and physical manifestations.

**Results:** Two-hundred and 86 respondents completed the survey. Thirty percent were unsatisfied with receiving genetic diagnoses. Satisfaction was predicted if communication was by geneticists (*p* = 0.004); provided face-to-face (*p* = 0.003); clearly explained (*p* < 0.001); and accompanied by support (*p* = 0.017). Parents obtained psychiatric information from non-professional sources more often than developmental (ϕ 0.26, *p* < 0.001) and physical manifestations (ϕ 0.21, *p* = 0.003), which mostly came from health professionals. Information from support organisations was more helpful than from geneticists (odds ratio [OR] 21.0, 95% CI 5.1 – 86.8, *p* < 0.001); paediatricians (OR 11.0, 1.4 – 85.2, *p* = 0.004); and internet sites (OR 15.5, 3.7 – 64.8, *p* < 0.001).

**Conclusion:** A paucity of professional information about psychiatric risks after genetic diagnosis may impede early diagnosis and intervention for children with high genotypic risks. Planned integration of genomic testing into mainstream services should include genetic counselling training to address the full spectrum of developmental, physical and psychiatric manifestations and timely provision of high-quality information.

## INTRODUCTION

Approximately 1 in 25 children in high income countries have neurodevelopmental disabilities (ND).[1] Clinical evaluation may include genetic investigations, which for many individuals can improve diagnostic accuracy, guide appropriate interventions and enhance genetic counselling for the family. Technological barriers to identifying genetic causes in severely disabled children have substantially diminished since the mid 2000s with the introduction of genome-wide diagnostic tests, primarily chromosomal microarray (CMA) assays capable of detecting sub-microscopic genomic imbalances known as Copy Number Variants (CNVs). Numerous genomic variants including CNVs and single nucleotide variants (SNVs) have been associated with ND and co-occurring psychiatric phenotypes.[**2–4**] For example, the 3Mb CNV diagnosed in 22q11.2 Deletion Syndrome (formerly Velocardiofacial Syndrome and DiGeorge syndrome) is causally associated with high rates of autism spectrum disorder (ASD), ADHD, schizophrenia, oppositional defiant disorders and anxiety.[**5–7**) Collectively, ND associated CNVs are implicated in up to 15% of children with severe developmental disorders who access genomic medicine services.[[8] Estimates suggest up to 42% of undiagnosed developmental disorders (DD) are due to *de novo* mutations in coding sequences about which risks for psychiatric complications are less well understood.[[9]

Variation in the penetrance of genomic variants associated with ND,[10–12] suggests health outcomes are susceptible to socio-environmental modification, additional undetected mutations and polygenic variation, presenting significant barriers to personalisation of medicine, genetic counselling and family-oriented information. However, genomic diagnosis is recognised as a major advantage to individuals with rare developmental disorders.[**13,14**] Progress towards introducing increasingly sophisticated whole genome and exome sequencing into clinical services is, however, placing additional demands on practitioners in terms of additional training requirements, clinic time, informed consent, and information provision.[**15–17**] The complexity of neurodevelopmental, physical and psychiatric outcomes associated with deleterious genomic variants presents important challenges for practitioners confronted by parents requiring detailed information about their child’s future health and wellbeing.[**18–20**]

The design and implementation of genomics medicine services for a broader range of neurodevelopmental conditions should account for the views and opinions of service users and recognise the need for relevant information both before and after genetic investigations. In this study we sought to address these issues by, (i) identifying key factors influencing parental satisfaction with genetics services, specifically the communication of diagnostic variants by clinical specialists; (ii) exploring how parents gain knowledge of developmental, psychiatric and physical manifestations of genomic variants; and (iii) comparing the availability, content and helpfulness of patient information obtained from health professionals, internet sites, voluntary support groups and other sources.

## METHODS

### Survey respondents and procedures

We designed a 46 item online survey, using Online Surveys (https://www.onlinesurveys.ac.uk, Jisc, Bristol UK), for parents of children aged 0-17 years with developmental, intellectual and congenital disorders having a clinical genetic diagnosis. The survey comprised 4 main sections: (1) family demographics, child’s genotype, reported medical, developmental and psychiatric diagnoses, family genetic history; (2) awareness of manifestations associated with child’s genetic diagnoses and sources of information to understand them; (3) ratings of the quantity, content and helpfulness of information from; and (4) experiences of services and receiving genetic test results. An introductory section included information about the research team, the purpose of the study, instructions for participating, participant confidentiality and data protection. Participants provided consent by agreeing with the statement, “I have read the information above and on the previous page, understand that my participation is voluntary, and I am happy to complete the questionnaire”. The study received ethical approval from Cardiff University School of Medicine Ethics Committee on 19 September 2014 (reference SMREC 14/34). Invitations to take part were distributed by advertisements in newsletters, websites and Facebook pages and by word-of-mouth at family support days sponsored by rare disorders support groups, including Unique – Understanding Rare Chromosome and Gene Disorders, Max Appeal, and Microdeletion 16p11.2 Support and Information UK.

### Statistical analyses

Response data was coded and downloaded from the survey website into the SPSS package (IBM Corp. SPSS Statistics for Mac, version 25.0, Armonk, NY, 2017) for statistical analysis. For identification of factors which influence parental satisfaction with genetics services we did logistic regression. We designed a hierarchical model including variables covering, (i) demographic data; (ii) components of genetic counselling process; and (iii) health providers involved in communicating results. The binary outcome variable was defined as parental satisfaction (or dissatisfaction) with the communication of diagnostic genetic test results by their health provider. The initial regression model incorporated family and child specific covariate using method ‘Enter’. Sequential hierarchical models incorporated genetic counselling and communication modality specific covariates. Where appropriate, ordinal variables were collapsed into fewer categories where responses were low. Odds ratios correspond to the exponentiated unstandardised coefficients (beta weights) for each variable. We employed a variety of tests to explore (a) sources of and (b) relative merits of information received by families after genetic testing. To compare information sources reported we used chi-square analyses of 2×2 contingency tables to examine relationships between 4 categories of manifestation, (i) developmental, (ii) physical, (iii) neuropsychiatric and (iv) other psychiatric disorders. Effect sizes were described in terms of Cramer’s phi coefficients (ϕ). Pairwise comparisons of information sources between individual manifestations was done by McNemar’s chi-square tests for marginal homogeneity in matched-pair binomial data. Odds ratios were calculated with VassarStats at http://www.vassarstats.net. To explore the relative quantity and quality of data from different sources we employed Wilcoxon’s signed-rank test to compare Likert scale responses and derived effect sizes (r) from Z scores.

## RESULTS

### Respondents

Two-hundred and eighty-six survey responses were recorded between 12 December 2014 and 31 May 2017. 199 respondents (70%) were located in the UK and 87 (30%) in the USA. Table 1 shows details of the respondents and their children. The most frequent reasons for referral to genetics services were developmental delay (N= 98, 34.4%), congenital anomaly (12.2%) and dysmorphic features (N=21, 7.3%). In addition to the referral indication for genetic testing, parents reported their children had multiple developmental, physical, neuropsychiatric and other mental health manifestations, a mean of 5.7 diagnoses. In terms of genetic diagnoses, 243 (85%) were diagnosed with chromosomal copy number variants (CNVs) which reportedly explained the referral. Of the CNVs reported, 116 (40.6%) were loci significantly associated with neurodevelopmental and psychiatric outcomes. Sixteen (5.6%) children had more than 1 CNV and 22 (7.7%) were diagnosed with single nucleotide variants (SNV) of which 5 were eponymous genetic syndromes. Thirty-nine parents (13.6%) reported their child’s CNV was inherited.

**Table 1.** Characteristics of the study sample. *1* Developmental Delay, Learning Disability, Speech and Language Delay. *2* Palatal, Cardiac, Respiratory, Musculoskeletal, Growth, Seizures/epilepsy, Sight, Hearing, Skin. *3* Attention deficit hyperactivity disorder, ADHD (N= 47), autism spectrum disorder, ASD (N= 86), obsessive compulsive disorder, OCD (N = 48), developmental coordination disorder, DCD (N = 49), dyslexia (N = 19). *4* Anxiety, Depression, Schizophrenia or psychosis. *5* Neurodevelopmental CNVs: 1q21.1 deletion, 1q21.1 duplication, 2p16.3 deletion (*NRXN1*), 9q34.3 deletion, 15q11.2 deletion, 15q11.2 duplication, 15q11.2 (NOS), 15q13.3 deletion, 15q13.3 duplication, 15q13.3 (NOS), 16p11.2 deletion, 16p11.2 duplication, 16p11.2 (NOS), 16p12.2 deletion, 16p13.11 deletion, 16p13.11 duplication, 16p13.11 (NOS), 17q12 deletion, 22q11.2 deletion, 22q11.2 distal deletion, 22q11.2 duplication.

| Respondents |  |  | Children |  |  | Parent reported diagnoses |  |  |
| --- | --- | --- | --- | --- | --- | --- | --- | --- |
| Location | N | % | Age (years) | Mean | Range | Clinical | N | Mean |
| UK | 199 | 69.9 | Age when diagnosed | 3.7 | 0-17 | Developmental<sub>1</sub> | 695 | 2.4 |
| USA | 87 | 30.4 | At time of survey | 8.5 | 0-42 | Physical<sub>2</sub> | 638 | 2.2 |
| Gender | N | % | Gender | N | % | Neuropsychiatric<sub>3</sub> | 230 | 0.8 |
| Female | 270 | 94.4 | Female | 125 | 43.7 | Mood and Psychotic<sub>4</sub> | 80 | 0.3 |
| Male | 16 | 5.6 | Male | 161 | 56.3 | **Total** | **1643** | **5.7** |

**Table 1.** Characteristics of the study sample.
| Relationship | N | % | Reason for referral | N | % | Genetic data | N | % |
| --- | --- | --- | --- | --- | --- | --- | --- | --- |
| Biological parent | 267 | 93.3 | Developmental delay | 98 | 34.3 | Neurodevelopmental CNV<sup>5</sup> | 116 | 40.6 |
| Adoptive parent | 10 | 3.5 | Congenital anomaly | 35 | 12.2 | Other CNV | 127 | 44.4 |
| Legal guardian | 2 | 0.7 | Dysmorphic features | 21 | 7.3 | Multiple CNVs | 16 | 5.6 |
| Other | 7 | 2.4 | Growth or stature | 16 | 5.6 | Named gene | 17 | 5.9 |
| Occupation | N | % | Neurological | 16 | 5.6 | Named syndrome | 5 | 1.7 |
| Employed | 141 | 49.3 | Multiple concerns | 13 | 4.5 | Unspecified | 5 | 1.7 |
| Carer | 78 | 27.3 | Learning disability | 11 | 3.8 | **Total** | 286 | 100 |
| Home maker | 58 | 20.3 | Family history | 10 | 3.5 | Inherited CNV | 39 | 13.6 |
| Other | 9 | 3.1 | Neurodevelopmental | 10 | 3.5 | Siblings with CNV | 20 | 7.0 |
|  |  |  | Neonatal concerns | 9 | 3.1 | Other relatives with CNV | 13 | 4.5 |
|  |  |  | Foetal anomaly | 7 | 2.4 |  |  |  |
|  |  |  | Metabolic | 2 | 0.7 |  |  |  |
|  |  |  | Unspecified | 38 | 13.3 |  |  |  |

### Reporting genetic diagnoses for developmental disorders

The great majority of respondents received their child’s genetic test result from paediatricians (130/286, 45.5%), clinical geneticists (116/286, 40.6%) or genetic counsellors (28/286, 9.8%). Parents were more likely to receive results from paediatricians than genetic specialists in the UK (107 vs 81) compared to those in the USA (22 vs 63, p <0.001). Eighty-five of 286 respondents (29.7%) were dissatisfied with how their child’s test result was communicated. Parents in the UK were more likely to be dissatisfied than in the USA (66/199 *vs* 19/87; *p* = 0.05). In terms of the specialism of health professionals delivering results, parents significantly were more likely to be satisfied when genetic specialists delivered results (116/144, 80.6%) as compared to paediatricians (78/129, 60.5%; *p* <0.001), which was accounted for by UK responses (figure 1 upper panel). More than one-in-three (101/286, 35.3%) were not satisfied with the explanation of their child’s genetic findings. Dissatisfaction was more prevalent among UK than USA respondents (78/199 *vs* 23/87, *p* = 0.038). Again parents were more likely to be satisfied by explanations given by genetic specialists than by paediatricians (106/144 *vs* 70/129, *p* = 0.001), which was also accounted for by UK responses (figure 1 lower panel). Around 3-in-10 parents (90/286, 31.5%) did not receive supporting information to accompany test results, which was more prevalent among UK respondents (36.2% *vs* 20.7%, *p* = 0.009). Almost three quarters of parents (208/286, 72.7%) reported not receiving support when test results were given, half (142/286, 49.7%) were not offered follow-up appointments.

**Figure 1.**
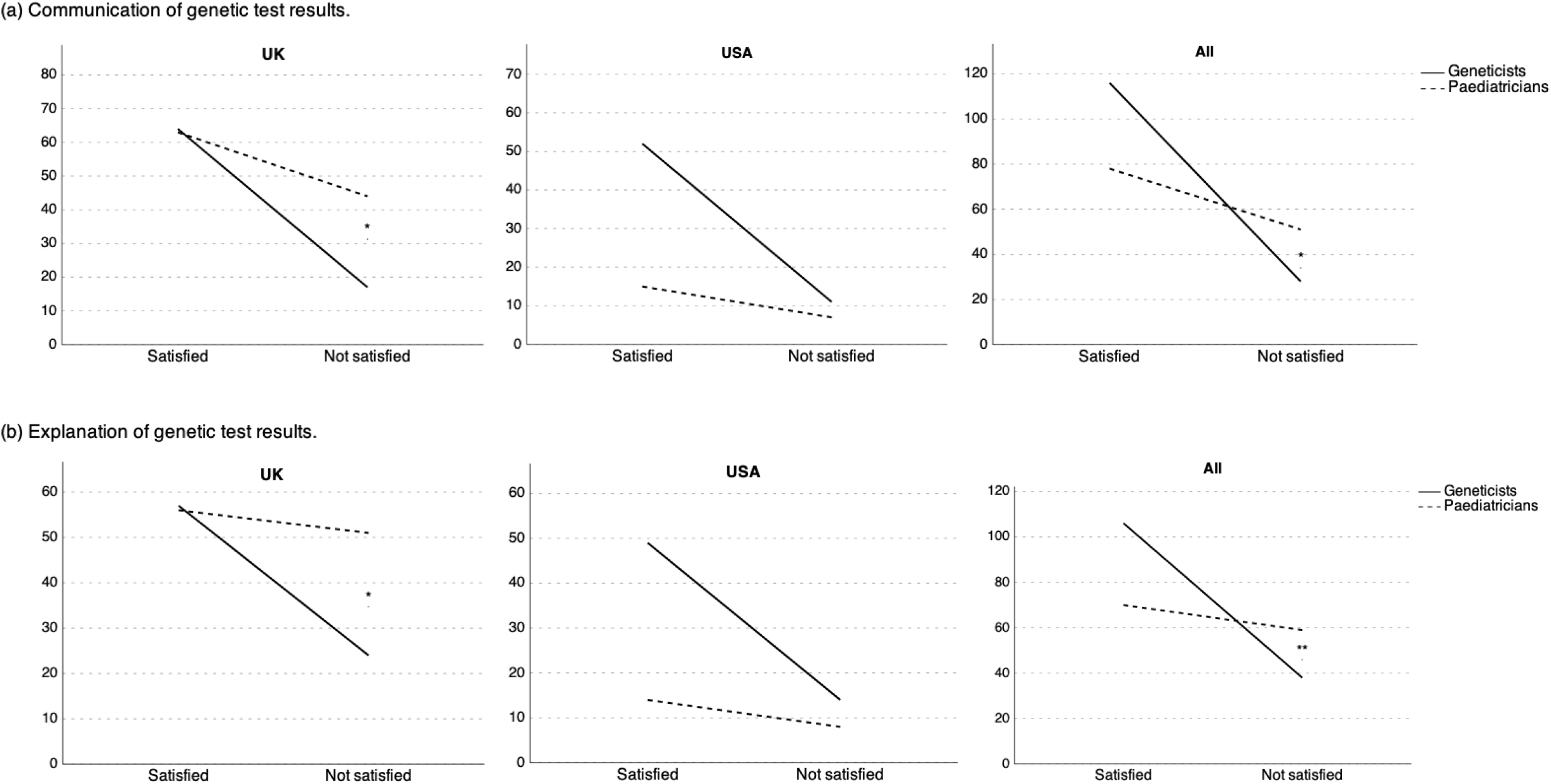
Comparative satisfaction with the delivery of genetic test results by genetic and paediatric specialists. Upper panel (a): Parents’ satisfaction with how genetic test results were communicated. Lower panel (b): Parents’ satisfaction with how genetic test results were explained. UK (N = 188); USA (N = 85); All (N = 273). Chi-Square test for independence: * *p* < 0.01; ** *p* < 0.001.

We used logistic regression to identify predictive factors for satisfaction with receiving genetic test results, incorporating 3 categories of variables; (i) child and family specific; (ii) genetic counselling; and (iii) modes of communication. A hierarchical model revealed satisfaction was most likely if, (i) results were presented by genetic specialists rather than paediatricians (OR = 2.97, CI 1.41–6.26); (ii) results were communicated in person instead of letter or telephone call (OR = 2.91, CI 1.41–6.26); (iii) results were satisfactorily explained (OR = 5.14, 95% CI 2.58–10.26); and (iv) support was given at the same time (OR = 2.99 CI 1.21– 7.36). Interestingly, parents of children identified as male were substantially more likely to be satisfied than those with females (final model OR = 2.56, CI 1.28–5.14). Receiving supplementary information from the practitioner or a follow-up appointment did not predict satisfaction. The final model accounted for 40.5% of the variance in the outcome variable (see Table in supplementary information).

### Sources of information on genomic disorders

We asked parents to indicate which sources they had used to gather information concerning manifestations associated with their child’s genetic variant, (i) health professionals; (ii) internet sites; (iii) voluntary sector support groups; and (iv) other lay sources (friends and family, books, leaflets and other materials). We compared sources used between 4 groups of manifestations, (i) developmental disorders; (ii) physical anomalies; (iii) neuropsychiatric disorders; and (iv) mood and psychotic disorders (defined in table 2). Information on developmental disorders was more likely to be given by health professionals at the time of genetic diagnosis or at follow-up compared with neuropsychiatric disorders (60.7% *vs* 37.9%), for which a majority parents used alternative sources (Cramer’s ϕ 0.22, *p* < 0.001). The disparity increased when comparing developmental disorders with mood and psychotic manifestations (60.7% *vs* 28.6% used health professional sources, (ϕ 0.29, *p* < 0.001) but not physical anomalies (table 2). Overall, fewer parents were informed about mental health manifestations 34.7% than developmental and physical manifestations (59%) by their clinician after genetic diagnosis as opposed to finding information from other sources, (ϕ 0.24, *p* <0.001). Following genetic diagnosis, fewer than 1-in-3 parents received information from their child’s Clinician about the possibility of mood and psychotic disorders (table 2).

**Table 2.** Sources of information reported by parents concerning manifestations of their child’s genetic diagnosis. Totals for each group of manifestations comprise cumulative frequencies for individual conditions: (1) Developmental = global developmental delay (N=274), intellectual disability (N=267), speech and language delay (N=257); (2) Physical = cardiac defects (N=153), palatal defects (N=97), seizures/epilepsy (N=166); (3) Neuropsychiatric = autism spectrum disorder (N=203), attention deficit hyperactivity disorder (N=130), obsessive compulsive disorder (N=140), developmental co-ordination disorder (N=115); (4) Mood and psychotic = anxiety (N=154), depression (N=85), schizophrenia or psychosis (N=73). Figures in parenthesis are counts for parent-reported associations between their child’s genotype and individual manifestations. *1* percentage values are for cumulative counts for individual manifestations in each group.

| Information source | Developmental and physical manifestations |  |  |  |  |  | Mental health manifestations |  |  |  |  |  |
| --- | --- | --- | --- | --- | --- | --- | --- | --- | --- | --- | --- | --- |
|  | Developmental |  | Physical |  | Combined |  | Neuropsychiatric |  | Mood + psychotic |  | Combined |  |
|  | N | %<sup>1</sup> | N | %<sup>1</sup> | N | %<sup>1</sup> | N | %<sup>1</sup> | N | %<sup>1</sup> | N | %<sup>1</sup> |
| **Health professional** |  |  |  |  |  |  |  |  |  |  |  |  |
| With diagnosis | 318 | 39.8 | 149 | 35.2 | 467 | 38.2 | 121 | 20.6 | 45 | 14.2 | 166 | 18.3 |
| At follow-up | 70 | 8.8 | 38 | 9.0 | 108 | 8.8 | 38 | 6.5 | 12 | 3.8 | 50 | 5.5 |
| Other | 96 | 12.0 | 49 | 11.6 | 145 | 11.9 | 64 | 10.9 | 34 | 10.7 | 98 | 10.8 |
| Total | 484 | 60.7 | 236 | 55.8 | 720 | 59.0 | 223 | 37.9 | 91 | 28.6 | 314 | 34.7 |
| **Non-professional** |  |  |  |  |  |  |  |  |  |  |  |  |
| Internet sites | 167 | 20.9 | 103 | 24.3 | 270 | 22.1 | 163 | 27.7 | 106 | 33.3 | 269 | 29.7 |
| Support groups | 74 | 9.3 | 40 | 9.5 | 114 | 9.3 | 100 | 17.0 | 66 | 20.8 | 166 | 18.3 |
| Other sources | 73 | 9.1 | 44 | 10.4 | 117 | 9.6 | 102 | 17.3 | 55 | 17.3 | 157 | 17.3 |
| Total | 314 | 39.3 | 187 | 44.2 | 501 | 41.0 | 365 | 62.1 | 227 | 71.4 | 592 | 65.3 |
| **Total observations** | **798** |  | **423** |  | **1221** |  | **588** |  | **318** |  | **906** |  |

The observation of increased psychiatric co-morbidity in individuals with neurodevelopmental CNVs (ND-CNVs) [**5**] prompted us to examine whether clinicians were more likely to offer information concerning risks for either neurodevelopmental or psychiatric outcomes to parents of children diagnosed with ND-CNVs (116/248, 46.8% reported CNVs) with, compared to those with other CNVs (132/248, 53.2%). For neurodevelopmental phenotypes (DCD, OCD, ADHD and ASD, defined in table 2), combined responses across all manifestations revealed parents of children with ND-CNVs were more likely to obtain information from lay sources than from their child’s clinician (Cramer’s ϕ 0.13, *p* = 0.004). The difference between CNV type was greatest for DCD (ϕ 0.29, *p* = 0.004). Similarly, for psychiatric disorders (schizophrenia or psychosis, anxiety and depression), in the event of receiving ND-CNV diagnoses, parents were more likely to employ lay sources of information (ϕ 0.16, *p* = 0.01). There were no differences between information sources for developmental or physical manifestations.

We then compared the main source of information (clinician delivering result *versus* all other sources) between individual manifestations parents associated with their child’s genetic diagnosis. Marginal homogeneity tests of matched pair data compared sources for each manifestation with reference phenotypes for 3 categories or clinical manifestation: ID (developmental disorders); cardiac anomalies (physical disorders); and ASD (psychiatric disorders). Compared to ID, information about all psychiatric manifestation was more likely to be obtained from sources other than clinicians. Odds ratios (OR) ranged from 2.7 for ASD (95% CI 1.5 – 4.8, *p* = 0.001) to 18.0 for depression (CI 2.4 – 134.8, *p* < 0.001) (figure 3, upper panel). Other than DD, there were no differences between the main source for ID and any developmental or physical disorder we tested (figure 2 upper panel). Comparisons with information sources for cardiac anomalies revealed a similar picture, with ORs ranging from 3.63 for ASD (CI 1.7 – 7.9, *p* = 0.001) to 31.0 for depression (CI 4.23 – 227.1, *p* < 0.001) (figure 2 middle panel). Finally, in comparison to ASD, information about anxiety, depression and OCD was more likely to be acquired from lay sources, OR range 6.0 for OCD (CI 1.8 – 20.4, *p* = 0.001) to 16.0 for anxiety (CI 2.1 – 120.7, *p* < 0.001) (figure 2 lower panel). Conversely, parents were more likely to be informed about cardiac anomalies, developmental delay, intellectual disability and speech and language delay by clinicians rather than obtaining information from other sources. Odds ratios varied between 2.7 for ID (CI 1.5 – 4.8, *p* = 0.001) and 12.8 for DD (CI 5.6 – 29.5, *p* < 0.001).

**Figure 2.**
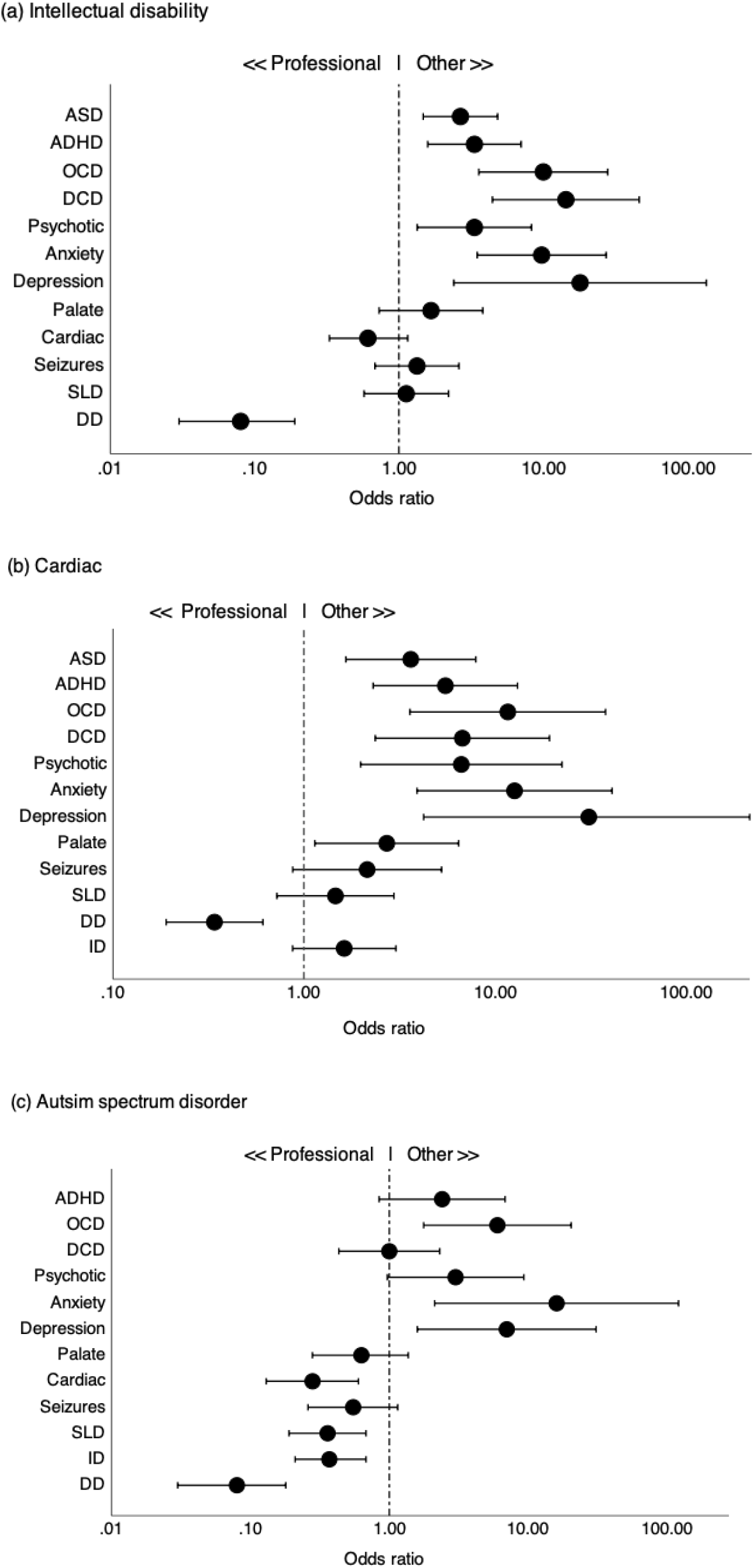
Variation in information sources for individual manifestations. Dot plots depicts odds ratios for pairwise comparisons between sources first used for information about recurrent CNV-associated manifestations compared with sources representative of three major classes of disorder: (a) developmental disorders (intellectual disability [ID]); (b) congenital disorders (cardiac defects); (c) neuropsychiatric disorders (autism spectrum disorder). Sources: (i) health professionals – clinician at time of genetic diagnosis; clinician at follow-up appointment; other health professional, (ii) other sources – internet sites; charity support organisations (including Facebook groups); friends and family; books and leaflets. Bars represent 95% confidence intervals. Manifestation groups: (i) developmental: developmental delay (DD); intellectual disability (ID); speech and language delay (SLD), (ii) physical: palatal defects; cardiac defects; seizures or epilepsy, (iii) neuropsychiatric: autism spectrum disorder (ASD); attention deficit hyperactivity disorder (ADHD); obsessive compulsive disorder (OCD); developmental coordination disorder (DCD), (iv) mood and psychotic: schizophrenia or psychosis (psychotic); anxiety; depression.

**Figure 3.**
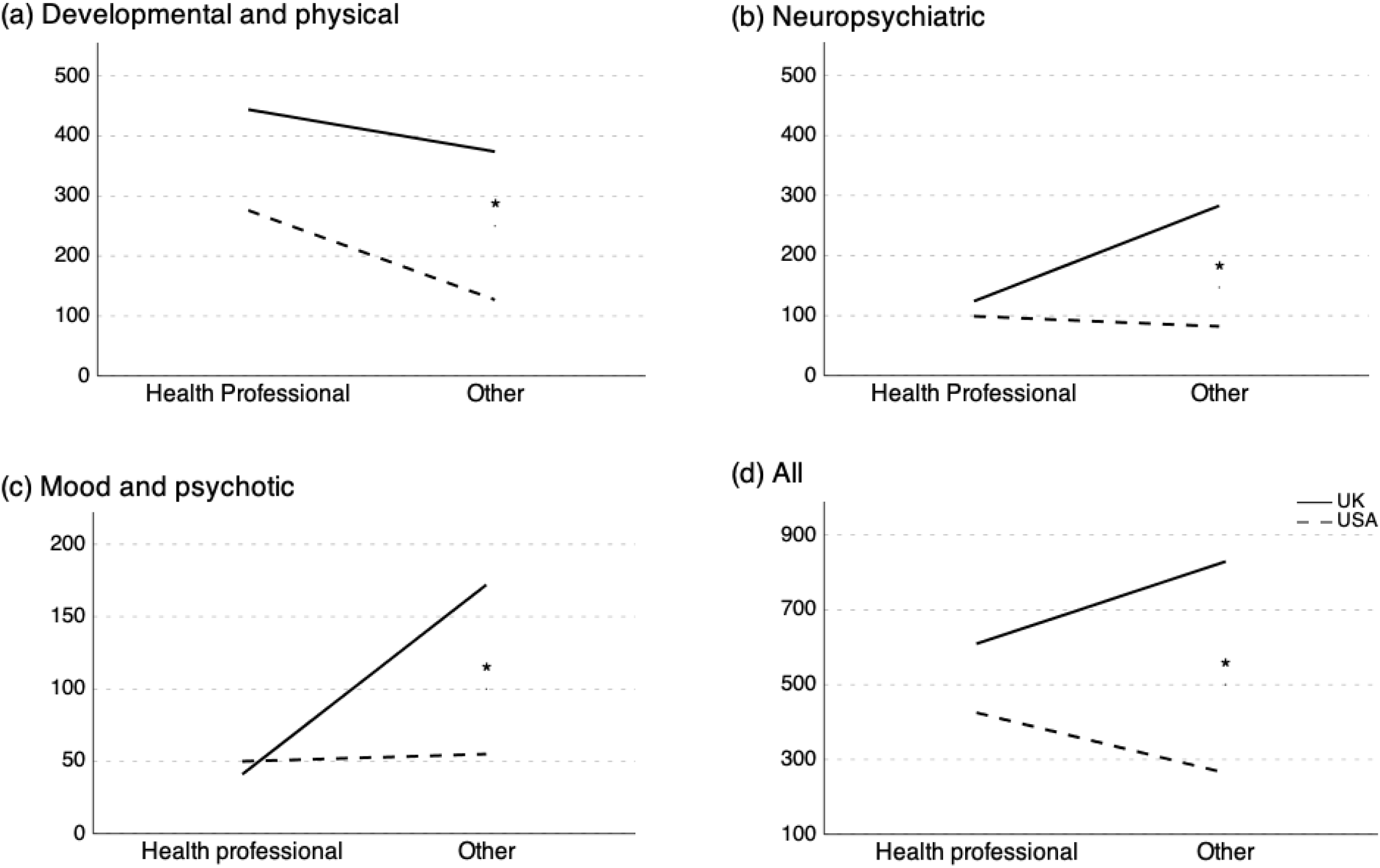
Comparisons of information sources concerning developmental, physical and psychiatric manifestations of genomic disorders reported by UK and USA respondents. Manifestations were grouped into (a) developmental and physical; (b) neuropsychiatric; (c) mood and psychotic; and (d) all combined (further defined in table 2). Information sources: (i) Health professional – clinician at time of genetic diagnosis, clinician at follow-up, or other health professional; (ii) Other – internet sites, charity support organisations (including Facebook groups), friends and family, books and leaflets. Chi-Square test for independence: * p < 0.001.

We explored whether large differences in the speciality of clinicians returning genetic test results in UK and USA associated with different patterns of information seeking by parents subsequent to diagnosis. Parents in the USA received more information on developmental and physical manifestations from health professionals (68.5% professional *vs* 31.5% other source) than in the UK (54.3% professional *vs* 45.7% other, Cramer’s ϕ 0.13, *p* <0.001) (see figure 3a). Between country differences were larger for psychiatric disorders. For neuropsychiatric manifestations (ASD, ADHD, OCD), 69.5% of UK parents obtained information from lay sources compared to 45.3% in the USA (ϕ 0.23, *p* < 0.001). For mood and psychotic disorders (anxiety, depression, schizophrenia, psychosis) less than 1-in-5 (19.2%) UK parents obtained information from clinicians, compared to 47.6% in the USA (ϕ 0.30, *p* <0.001, figure 3c). Overall, UK parents sought information alternative sources more often than their US counterparts (57.6% *vs* 38.3%, Cramer’s ϕ 0.18, *p* < 0.001, figure 3d).

Finally, we evaluated respondents’ opinions on the quantity, content and helpfulness of information obtained according to its source. Analysis of mean ranks revealed the amount of information available from support groups was more optimal than from paediatricians (r 0.47, *p* <0.001), geneticists (r 0.51, *p* <0.001) and internet sites (r 0.38, *p* <0.001) (table 3). Similarly, information from internet sites was more optimal than that from paediatricians (r 0.34, *p* <0.001) and geneticists (r 0.32, *p* <0.001). In terms of the quality of its content, information from support groups was more optimal than internet sites (r 0.31, *p* <0.001), geneticists (r 0.29, *p* <0.001) and paediatricians (r 0.14, *p* = 0.012). In terms of the usefulness of information, pairwise comparisons revealed that support groups strongly outperformed internet sites (Odds ratio 15.5, 95% CI 3.71–64.77, *p* <0.001), paediatricians (OR 11.0, 1.42–85.2, *p* = 0.004) and geneticists (OR 21.0, 5.08–86.75, *p* <0.001), Internet derived information was more helpful than content given by geneticists (OR 2.5, CI 1.45– 4.32, *p* = 0.001) but not paediatricians (table 3). Collectively, lay sources of information were consistently more helpful to parents than information given by health professionals (OR 2.2, CI 1.37–3.53, *p* = 0.001, table 3).

**Table 3.** Comparative value of information from health professionals and non-professional sources. *1* Amount of information (too little; sufficient; too much) and *2* Content of information (too complicated; clear and comprehendible; not relevant) was assessed by pairwise comparison of responses; *3* The helpfulness of information (helpful *vs* not helpful) was assessed by testing differences in paired proportions.

| Comparison | Observations | Amount of information<sup>1</sup> |  | Content of information<sup>2</sup> |  | Helpfulness of information<sup>3</sup> |  |
| --- | --- | --- | --- | --- | --- | --- | --- |
|  |  | *r* | *p* value | *r* | *p* value | Odds ratio (95% CI) | *p* value |
| Geneticists vs Paediatricians | 142 | 0.03 | 0.679 | 0.07 | 0.232 | 2.25 (0.99 – 5.17) | 0.05 |
| Internet sites vs Paediatricians | 182 | 0.34 | < 0.001 | 0 | 0.96 | 1.88 (0.80 – 4.42) | 0.144 |
| Support groups vs Paediatricians | 120 | 0.47 | < 0.001 | 0.14 | 0.012 | 11.0 (1.42 – 85.20) | 0.004 |
| Internet sites vs Geneticists | 360 | 0.32 | < 0.001 | 0.12 | 0.035 | 2.5 (1.45 – 4.32) | 0.001 |
| Support groups vs Geneticists | 252 | 0.51 | < 0.001 | 0.29 | < 0.001 | 21.0 (5.08 – 86.75) | < 0.001 |
| Support groups vs Internet sites | 330 | 0.38 | < 0.001 | 0.31 | < 0.001 | 15.5 (3.71 – 64.77) | < 0.001 |

## DISCUSSION

This is the first quantitative survey of its kind to investigate how parents experience the communication of genomic tests for developmental disorders and their endeavours to gain knowledge about health implications associated with their child’s diagnosis. At a time of rapid advances in diagnosing rare disorders and diversification of genetics services, this is an important topic. Our survey reveals parents have mixed experiences of attending genetics services. A third of those in the UK reported dissatisfaction with the communication of test results. Information provided by health specialists was less helpful and of inferior quality compared to information respondents obtained from other sources, particularly voluntary support groups. Information provided by health specialists was predominantly focused on developmental and physical challenges typically present at diagnosis and parents relied heavily on their own resources to find information about psychiatric manifestations associated with their child’s genetic variant. Families in the UK were more reliant than the USA on internet sites and voluntary sector organisations for information, particularly for neurodevelopmental and psychiatric disorders.

The findings extend previous evidence showing that families often have difficulties obtaining satisfactory information from clinical specialists to aid comprehension of genomic tests results.[**21–23**] We have revealed new evidence concerning a broad range of genomic disorders consistent with earlier findings concerning 22q11.2 deletion syndrome - one of the most frequently diagnosed genomic disorders - in which parents predominantly obtained psychiatric information from internet sites and support groups.[**24,25**] Consistent with studies of rare disorders more generally, we found that most respondents resorted to internet searches to help comprehend their child’s diagnosis.[**26**] However, many families do not seek medical information online, and this may particularly be the case for more socioeconomically deprived groups.[**27**] Limited access to information is associated with increased stress and uncertainty for parents and negatively influences engagement with healthcare services, potentially negating some the advantages of having precision diagnoses.[**28**] Our survey did not gather sufficient demographic data to determine whether hard-pressed families were less likely to seek online content. However, we are concerned that socially disadvantaged families may not have the same degree of access to high quality information, support and services to which they are entitled.[**29**] This is emphasised by our finding that, as in previous studies, parents often consider information from clinical specialists to be overcomplicated, irrelevant or unhelpful compared to online content or from support groups.[**30**]

Our study showed that paediatric and genetic specialists tend to focus on providing information about developmental and physical disabilities at the time of giving genetic test results, suggesting either that information about psychiatric risks may be less readily available, or that informing parents about risks of behavioural, emotional and psychotic disorders is less relevant when communicating test results.[**31–33**] In the context of communicating pathogenic neurodevelopmental CNVs with significant evidence of psychiatric, our findings indicated parents were no more likely to be informed about these than were others. Moreover, our findings suggest parents of children with ND-CNVs tend to find psychiatric information through alternative sources including internet searches, suggesting extensive evidence on neuropsychiatric genotype-phenotype associations in ND-CNVs is yet to be incorporated into genetic counselling for individuals with developmental disorders. Often cited concerns over overloading parents with information in the aftermath of a genetic diagnosis should be balanced against the best interests of children and parents’ desires for comprehensive health information.[**34–36**] We recommend the translation of evidence on adverse neuropsychiatric outcomes into care guidelines for ND-CNVs coupled with mechanisms ensuring timely provision of family-oriented information which accounts for the needs and wishes of families over time.[**37–39**]

Our findings revealed differences in the communication of test results and information gathering between UK and US families. More than half of UK parents received their child’s diagnosis from paediatricians, compared to one-in-five in the USA. Parents in the USA received a broader range of medical and mental health information from their clinician, whereas UK parents obtain most neurodevelopmental and psychiatric information from alternative sources. However, the lower frequency of provision of psychiatric information by clinicians the UK was not explained by the smaller proportion of parents receiving their result from genetic specialists. As such, our survey failed to uncover evidence explaining why provision of psychiatric information from clinical professionals is relatively infrequent the UK, or, conversely why parents in US receive a broader range of information from their doctor. Other factors may account for these differences, such as healthcare service models, availability and duration of face-to-face appointments, or differences in professional development and training.

Opinions vary on the benefits and risks of online health information, ranging from concerns among health professionals over accuracy, relevance and under-regulation, to endorsements from sociologists in support of its contribution to client empowerment. Although an understudied topic, evidence has been published that the general public adopt contingent behaviours towards online content, discriminating between trustworthy and untrustworthy content in order to supplement rather than replace traditional media.[**40**] Importantly, universal online search methods are increasingly concordant with the structure of internet health information and the hierarchical nature of results created by popular search engines. Therefore, we recommend initiatives which support clinicians to improve access to comprehensive, high quality information around the time of diagnosis, including signposting to digital content beyond traditional media and support voluntary support groups to innovate new ways of supporting children with complex disabilities.

The findings presented here are timely; paediatric genetics services need to strike a fine balance between delivering high quality services for escalating referrals and performing increasingly sophisticated tests with lengthy consent procedures. That a significant proportion of respondents in our survey expressed dissatisfaction receiving genetic diagnoses highlights the challenge facing specialised clinical services. As the availability of genomic testing in mainstream services increases, the demand for relevant patient-oriented information concerning genetic influences on psychiatric risks are predicted to increase.[**41,42**] However, informing patients and families about complex and uncertain implications of genetic tests is challenging, requiring specialised expertise covering multiple domains of physical, medical and mental health, sensitive risk counselling and facilitation of parents’ adaptation to the diagnosis. Personalising psychiatric risk across the lifespan such that families and patients understand the true nature of their risks is likely to become a priority for psychiatrists and other specialists working in neuropsychiatric disorders. Without a sound understanding of these risks, parents will be less well equipped to recognize and respond to symptoms and access early interventions which – especially in the context of psychotic disorders – can lead to better long term outcomes.[**43,44**] We recommend that education and training in brain disorders is prioritised for all clinical specialties considering genetic testing.[**45,46**] Curriculum content should be developed accordingly, ensuring genetic counselling includes meaningful conversations about the full spectrum of medical and mental health risks accompanied by contemporary, relevant information. Fortuitously, evidence revealing complex but broadly similar psychiatric outcomes for children with recurrent neurodevelopmental CNVs suggests integrating a coordinated general approach to psychiatric care planning into a dedicated multidisciplinary clinical pathway for such children could be justified, with individual tailoring for genotype-specific risks.[**47**] This would require fundamental changes to service configuration and delivery and greater awareness of this significant population among health providers and other professionals.

Our study has limitations. Recruitment was biased in favour of parents who engage with the support groups who promoted the survey, potentially limiting the generalisability of our findings. The survey was designed with broad accessibility in mind. However, online surveys require significant internet literacy and time for participation, which may be difficult for some families. Parents who readily access the internet, are, conceivably, more likely to use it for seeking information related to the content of this study. The survey was overwhelmingly completed by mothers. However, families with severely disabled children are rare and not representative of the general population and this may be a reflection of the socio-domestic influences on caregiving in the context of disability.[**48**] Also, we were unable to independently verify health information provided by parents. Self-report surveys have recognised shortcomings exacerbated by respondents potentially having to recall facts and experiences several years after the event. Despite these limitations, the views and experiences expressed by a large number of families provides a timely insight into the current state of medical health information, clinical services and lay support for neuro-disability communities.

In summary, our findings reveal that parents often feel inadequately informed by their clinical specialists about potential neurodevelopmental and psychiatric challenges in the context of paediatric genetic testing. Parents search extensively for information about their child’s genetic diagnosis, retrieving neuropsychiatric information primarily from internet sites and lay support groups, where accuracy and validity is unregulated and likely less reliable. We believe the current results are important in informing service development and training in both clinical genetics and psychiatry. In particular, they point to the need for closer integration of medical genetics and psychiatry to address the needs of those receiving a genetic diagnosis. Future initiatives should focus on identifying measures to promote the inclusion of psychiatric risk information, provide greater support in genetic counselling clinics and ensuring that education and training for practitioners encompass the full spectrum of neurodevelopmental and mental health challenges faced by children with genomic disorders.

## Data Availability

Will individual participant data be available? Yes
What data will be shared? All of the relevant participant responses to the survey, after deidentification
What other documents be available Yes, the survey questions
When will the data be available? Immediately after publication
With whom? Researchers who provide a reasonable request based on a methodological sound proposal
For what types of analysis? To achieve the aims of their submitted proposal
How will the data be made available? In response to email sent to the corresponding author (ORCID iD 0000-0002-4426-3254).
A link to the survey data (after deidentification) will be provided to approved requests. Proposals may be submitted up to 36 months following the article’s publication.

## ACKNOWLEDGEMENTS

We are grateful to all the families who helped with this study. We thank Dr Beverley Searle and all the staff at the charity Unique - Understanding Rare Chromosome and Gene Disorders, for their support and for promoting the study. We also wish to thank Max Appeal for their assistance with the advertising the survey. We are grateful to Dr Michael Arribas Ayllon (School of Social Sciences, Cardiff University) for his valuable contribution. The work was supported by The Waterloo Foundation (Grant No. 506926) and the Medical Research Council (Grant No. MR/N022572/1). The funders had no role in the study design, data collection, analysis and interpretation of findings and made no contribution to writing the report. All authors had full access to all the data collected in the survey. The corresponding author had final responsibility for the decision to submit for publication.

## REFERENCES

1. Emerson E. Deprivation, ethnicity and the prevalence of intellectual and developmental disabilities. J Epidemiol Community Health 2012;66:218–24.

2. Malhotra D, Sebat J. CNVs: harbingers of a rare variant revolution in psychiatric genetics. Cell 2012;148:1223–41.

3. Cooper GM, Coe BP, Girirajan S, Rosenfeld JA, Vu TH, Baker C, Williams C, Stalker H, Hamid R, Hannig V, Abdel-Hamid H, Bader P, McCracken E, Niyazov D, Leppig K, Thiese H, Hummel M, Alexander N, Gorski J, Kussmann J, Shashi V, Johnson K, Rehder C, Ballif BC, Shaffer LG, Eichler EE. A copy number variation morbidity map of developmental delay. Nat Genet 2011;43:838–46.

4. Singh T, Walters JTR, Johnstone M, Curtis D, Suvisaari J, Torniainen M, Rees E, Iyegbe C, Blackwood D, McIntosh AM, Kirov G, Geschwind D, Murray RM, Di Forti M, Bramon E, Gandal M, Hultman CM, Sklar P, Palotie A, Sullivan PF, O’Donovan MC, Owen MJ, Barrett JC. The contribution of rare variants to risk of schizophrenia in individuals with and without intellectual disability. Nat Genet 2017;49:1167–73.

5. Schneider M, Debbané M, Bassett AS, Chow EWC, Fung WLA, Marianne B.M. van den Bree MO, Murphy KC, Niarchou M, Kates WR, Antshel KM, Fremont W, McDonald-McGinn DM, Gur RE, Zackai EH, Vorstman J, Duijff SN, Klaassen PWJ, Swillen A, Gothelf D, Green T, Weizman A, Amelsvoort T Van, Evers L, Boot E, Shashi V, Hooper SR, Bearden CE, Jalbrzikowski M, Marco Armando, Stefano Vicari DGM, Ousley O, Campbell LE, Simon TJ, Stephan Eliez and for the IC on B and B in 22q11.. DS. Psychiatric Disorders From Childhood to Adulthood in 22q11.2 Deletion Syndrome: Results From the International Consortium on Brain and Behavior in 22q11.2 Deletion Syndrome. Am J Psychiatry 2014;29:997–1003.

6. Monks S, Niarchou M, Davies AR, Walters JTR, Williams N, Owen MJ, van den Bree MBM, Murphy KC. Further evidence for high rates of schizophrenia in 22q11.2 deletion syndrome. Schizophr Res 2014;153:231–6.

7. Niarchou M, Zammit S, van Goozen SHM, Thapar A, Tierling HM, Owen MJ, van den Bree MBM. Psychopathology and cognition in children with 22q11.2 deletion syndrome. Br J Psychiatry 2014:46–54.

8. Miller DT, Adam MP, Aradhya S, Biesecker LG, Brothman AR, Carter NP, Church DM, Crolla J a., Eichler EE, Epstein CJ, Faucett WA, Feuk L, Friedman JM, Hamosh A, Jackson L, Kaminsky EB, Kok K, Krantz ID, Kuhn RM, Lee C, Ostell JM, Rosenberg C, Scherer SW, Spinner NB, Stavropoulos DJ, Tepperberg JH, Thorland EC, Vermeesch JR, Waggoner DJ, Watson MS, Martin CL, Ledbetter DH. Consensus Statement: Chromosomal Microarray Is a First-Tier Clinical Diagnostic Test for Individuals with Developmental Disabilities or Congenital Anomalies. Am J Hum Genet 2010;86:749–64.

9. Deciphering Developmental Disorders Study. Prevalence and architecture of de novo mutations in developmental disorders. Nature 2017;542:433–7.

10. Kirov G, Rees E, Walters JTR, Escott-Price V, Georgieva L, Richards AL, Chambert KD, Davies G, Legge SE, Moran JL, McCarroll S a, O’Donovan MC, Owen MJ. The Penetrance of Copy Number Variations for Schizophrenia and Developmental Delay. Biol Psychiatry 2013;75:378–85.

11. Rees E, Kendall K, Pardiñas AF, Legge SE, Pocklington A, Escott-price V, Maccabe JH, Collier DA, Holmans P, Donovan MCO, Owen MJ, Walters JTR, Kirov G. Analysis of Intellectual Disability Copy Number Variants for Association With Schizophrenia. JAMA Psychiatry 2016;73:1–7.

12. Owen MJ, O’Donovan MC, Thapar A, Craddock N. Neurodevelopmental hypothesis of schizophrenia. Br J Psychiatry 2011;198(3):173–5.

13. Bouwkamp CG, Kievit AJA, Markx S, Friedman JI, Van Zutven L, Van Minkelen R, Vrijenhoek T, Xu B, Sterrenburg-Van De Nieuwegiessen I, Veltman JA, Bonifati V, Kushner SA. Copy number variation in syndromic forms of psychiatric illness: The emerging value of clinical genetic testing in psychiatry. Am J Psychiatry 2017;174:1036–50.

14. Wright CF, Fitzgerald TW, Jones WD, Clayton S, McRae JF, Van Kogelenberg M, King DA, Ambridge K, Barrett DM, Bayzetinova T, Bevan AP, Bragin E, Chatzimichali EA, Gribble S, Jones P, Krishnappa N, Mason LE, Miller R, Morley KI, Parthiban V, Prigmore E, Rajan D, Sifrim A, Swaminathan GJ, Tivey AR, Middleton A, Parker M, Carter NP, Barrett JC, Hurles ME, Fitzpatrick DR, Firth H V. Genetic diagnosis of developmental disorders in the DDD study: A scalable analysis of genome-wide research data. Lancet 2015;385:1305–14.

15. Slade I, Subramanian DN, Burton H. Genomics education for medical professionals - the current UK landscape. Clin Med 2016;16:347–52.

16. Burton H, Cole T, Lucassen AM. Genomic medicine?: challenges and opportunities for physicians The impact of genomic technologies on diagnosis. Clin Med 2012;12:416– 9.

17. Dheensa S, Lucassen A, Fenwick A. Fostering trust in healthcare: Participants’ experiences, views, and concerns about the 100,000 genomes project. Eur J Med Genet 2018;62:335–41.

18. Limb L, Nutt S. Improving Lives, Optimising Resources?: A Vision for the UK Rare Disease Strategy. 2011. Available from https://www.raredisease.org.uk/wp-content/uploads/sites/7/2018/04/improving-lives-optmising-resources-a-vision-for-the-uk-rare-disease-strategy.pdf.

19. Wright CF, FitzPatrick DR, Firth H V. Paediatric genomics: Diagnosing rare disease in children. Nat Rev Genet 2018;19:253–68.

20. Ryten M, Owen M, Kirov G, Cuthbert A, Saklavata J, Simpson M, Treacy B, Mohammed S. Provision of clinical genetic tests within the NHS associated with Mental Health Disorders. UK Genetic Testing Network Brief Report 2016. Available from: https://ukgtn.nhs.uk/fileadmin/uploads/ukgtn/Documents/Resources/Library/Reports_Guidelines/Dec-Mental_Health_Briefing_Report_FINAL.pdf

21. Ashtiani S, Makela N, Carrion P, Austin J. Parents’ experiences of receiving their child’s genetic diagnosis: A qualitative study to inform clinical genetics practice. Am J Med Genet 2014;164:1496–502.

22. Maya I, Sharony R, Yacobson S, Kahana S, Yeshaya J, Tenne T, Agmon-Fishman I, Cohen-Vig L, Goldberg Y, Berger R, Basel-Salmon L, Shohat M. When genotype is not predictive of phenotype: Implications for genetic counseling based on 21,594 chromosomal microarray analysis examinations. Genet Med 2018;20:128–31.

23. Reiff M, Bernhardt BA, Mulchandani S, Soucier D, Cornell D, Pyeritz RE, Spinner NB. “What does it mean?”: Uncertainties in understanding results of chromosomal microarray testing. Genet Med 2012;14:250–8.

24. Hercher L, Bruenner G. Living with a child at risk for psychotic illness: The experience of parents coping with 22q11 deletion syndrome: An exploratory study. Am J Med Genet 2008;146:2355–60.

25. van den Bree MBM, Miller G, Mansell E, Thapar A, Flinter F, Owen MJ. The internet is parents’ main source of information about psychiatric manifestations of 22q11.2 deletion syndrome (22q11.2DS). Eur J Med Genet 2013;56:439–41.

26. Nicholl H, Tracey C, Begley T, King C, Lynch AM. Internet Use by Parents of Children With Rare Conditions: Findings From a Study on Parents’ Web Information Needs. Eysenbach G, editor. J Med Internet Res 2017;19:e51.

27. Roche MI, Skinner D. How parents search, interpret, and evaluate genetic information obtained from the internet. J Genet Couns 2009;18:119–29.

28. Goodwin J, Schoch K, Shashi V, Hooper SR, Morad O, Zalevsky M, Gothelf D, Campbell LE. A tale worth telling: the impact of the diagnosis experience on disclosure of genetic disorders. J Intellect Disabil Res 2014;59:474–86.

29. Ramsey I, Corsini N, Peters MDJ, Eckert M. A rapid review of consumer health information needs and preferences. Patient Educ Couns 2017;100:1634–42.

30. Gilmore L. Supporting families of children with rare and unique chromosome disorders. Res Pract Intellect Dev Disabil 2018;5:8–16.

31. Baughman ST, Morris E, Jensen K, Austin J. Disclosure of psychiatric manifestations of 22q11.2 deletion syndrome in medical genetics: A 12-year retrospective chart review. Am J Med Genet 2015;167:2650–56.

32. Martin M, Mikhaelian M, Cytrynbaum, Shuman C, Chitayat DA, Weksberg R. 22q11.2 Deletion Syndrome: Attitudes towards Disclosing the Risk of Psychiatric Illness. J Genet Couns 2012;21:825–34.

33. Morris E, Inglis A, Friedman J, Austin J. Discussing the psychiatric manifestations of 22q11.2 deletion syndrome: An exploration of clinical practice among medical geneticists. Genet Med 2013;15:713–20.

34. Walser SA, Kellom KS, Palmer SC, Bernhardt BA. Comparing genetic counselor’s and patient’s perceptions of needs in prenatal chromosomal microarray testing. Prenat Diagn 2015;35:870–8.

35. Dondorp W, Sikkema-Raddatz B, de Die-Smulders C, de Wert G. Arrays in postnatal and prenatal diagnosis: An exploration of the ethics of consent. Hum Mutat 2012;33:916–22.

36. Merrill SL, Guthrie KJ. Is it Time for Genomic Counseling? Retrofitting Genetic Counseling for the Era of Genomic Medicine. Curr Genet Med Rep 2015;3:57–64.

37. Hart SJ, Schoch K, Shashi V, Callanan N. Communication of Psychiatric Risk in 22q11.2 Deletion Syndrome: A Pilot Project. J Genet Couns 2016;25:6–17.

38. Costain G, Chow EWC, Ray PN, Bassett AS. Caregiver and adult patient perspectives on the importance of a diagnosis of 22q11.2 deletion syndrome. J Intellect Disabil Res 2012;56:641–51.

39. Bassett AS, McDonald-McGinn DM, Devriendt K, Digilio MC, Goldenberg P, Habel A, Marino B, Oskarsdottir S, Philip N, Sullivan K, Swillen A, Vorstman J, Anne S. Bassett, Donna M. McDonald-McGinn, Koen Devriendt, Maria Cristina Digilio, Paula Goldenberg, MSW, Alex Habel, Bruno Marino, Solveig Oskarsdottir, Nicole Philip, Kathleen Sullivan, Ann Swillen JV, Bassett AS, McDonald-McGinn DM, Devriendt K, Digilio MC, Goldenberg P, Habel A, Marino B, Oskarsdottir S, Philip N, Sullivan K, Swillen A, Vorstman J. Practical Guidelines for Managing Patients with 22q11.2 Deletion Syndrome. J Pediatr 2011;159:332–39.

40. Nettleton S, Burrows R, O’Malley L. The mundane realities of the everyday lay use of the internet for health, and their consequences for media convergence. Sociol Heal Illn 2005;27:972–92.

41. Wolfe K, Stueber K, McQuillin A, Jichi F, Patch C, Flinter F, Strydom A, Bass N. Genetic testing in intellectual disability psychiatry: Opinions and practices of UK child and intellectual disability psychiatrists. J Appl Res Intellect Disabil 2018;31:273–84.

42. Baker K, Costain G, Fung WLA, Bassett AS. Chromosomal microarray analysis - a routine clinical genetic test for patients with schizophrenia. Lancet Psychiatry 2014;1:329–31.

43. Onwumere J, Bebbington P, Kuipers E. Family interventions in early psychosis: specificity and effectiveness. Epidemiol Psychiatr Sci 2011;20:113–9.

44. Valmaggia R, K McGuire P, Fusar-Poli P, Howes O, McCrone P. Economic Impact of Early Detection and Early Intervention of Psychosis. Curr Pharm Des 2012;18:592–5.

45. O’Donovan MC, Owen MJ. The implications of the shared genetics of psychiatric disorders. Nat Med 2016;22:1214–19.

46. Hall J, Owen MJ. Psychiatric classification - A developmental perspective. Br J Psychiatry 2015;207:281–2.

47. Chawner SJRA, Owen MJ, Holmans P, Raymond FL, Skuse D, Hall J, van den Bree MBM. Genotype–phenotype associations in children with copy number variants associated with high neuropsychiatric risk in the UK (IMAGINE-ID): a case-control cohort study. Lancet Psychiatry 2019;6:493–505.

48. Sharma N, Chakrabarti S, Grover S. Gender differences in caregiving among family - caregivers of people with mental illnesses. World J Psychiatry 2016;6:7–17.

